# Impact of SARS-CoV-2 infection on vaccine-induced immune responses over time

**DOI:** 10.1101/2021.10.16.21264948

**Authors:** Sebastian Havervall, Ulrika Marking, Nina Greilert-Norin, Max Gordon, Henry Ng, Wanda Christ, Mia Phillipson, Peter Nilsson, Sophia Hober, Kim Blom, Jonas Klingström, Sara Mangsbo, Mikael Åberg, Charlotte Thålin

**Affiliations:** Department of Clinical Sciences, Karolinska Institutet Danderyd Hospital, Stockholm, Sweden; Department of Medical Cell Biology and SciLifeLab, Uppsala University, Uppsala, Sweden; Center for Infectious Medicine, Department of Medicine Huddinge, Karolinska Institutet, Stockholm, Sweden; Department of Protein Science, KTH Royal Institute of Technology, SciLifeLab, Stockholm, Sweden; Department of Pharmacy and SciLifeLab, Uppsala University, Uppsala, Sweden; Department of Medical Sciences, Clinical Chemistry and SciLifeLab, Uppsala University, Uppsala, Sweden

**Author notes:** Corresponding author. Mail. These authors contributed equally (Mikael Åberg, Charlotte Thålin).

## Abstract

People with previous SARS-CoV-2 infection mount potent immune responses to COVID-19 vaccination, but long-term effects of prior infection on these immune responses are unknown. We investigated the long-term impact of prior SARS-CoV-2 infection on humoral and cellular immune responses in healthcare workers receiving the mRNA BNT162b2 or the adenovirus vectored ChAdOx1 nCoV-19 vaccine. Vaccination with both vaccine platforms resulted in substantially enhanced T cell immune responses, antibody responses to spike and neutralizing antibodies effective against ten SARS-CoV-2 variants following SARS-CoV-2 infection, compared to in naïve individuals. The enhanced immune responses sustained over seven months following vaccination. These findings imply that prior infection should be taken into consideration when planning booster doses and design of current and future COVID-19 vaccine programs.

**One-Sentence Summary:** SARS-CoV-2 infection prior to vaccination leads to substantial and durable increases in immune memory responses.

---

Clinical trials and post marketing effectiveness data have shown that currently used COVID-19 vaccines protect strongly against hospitalization and death *(1-3)*. However, real-world efficacy estimates are affected by population demographics, characteristics of circulating SARS-CoV-2 variants, vaccine protocols and time since vaccination. An increased risk of breakthrough infections is now observed, partly explained by immune waning *(4-8)*, and third vaccine doses are therefore being administered. A robust immune response after infection or vaccination is based on the induction of memory B and T cells generating virus specific antibodies and T-cell responses *(9-13)*. Antibody levels have been shown to correlate inversely with risk of SARS-CoV-2 infection *(14, 15)* and may, with standardized readouts *(16)*, be used as a marker for correlates of protection. Time between prime and boost *(17)*, the number of boosters administrated as well as infection prior to vaccination *(18)* impact the breadth and duration of immune responses. As an increasing number of persons become infected globally, vaccination post SARS-CoV-2 infection will become more frequent. Prior SARS-CoV-2 infection has been reported to positively impact vaccine responses *(9, 19-24)* but little is known regarding long term effects. To optimize immunization programs, it is therefore of importance to study duration of immune responses including direct comparisons of vaccine platforms and the long-term effect of prior SARS-CoV-2 infection on subsequent vaccine-induced responses in real world evidence studies.

Using longitudinally collected blood samples from the COMMUNITY (COVID-19 Immunity) study *(13, 24-26)* we herein report binding and pseudo-neutralizing antibody titers and memory T cell responses elicited over seven months following mRNA BNT162b2 (Comirnaty, Pfizer BioNTech) and over three months following adenovirus-vectored ChAdOx1 nCoV-19 (Vaxzevria, AstraZeneca) vaccination in 517 healthcare workers (HCW) with and without confirmed SARS-CoV-2 infection prior to vaccination.

## Results

The COMMUNITY study enrolled 2149 HCW at Danderyd Hospital, Stockholm, Sweden, between April and May 2020. Starting January 2021 all HCW at Danderyd Hospital were offered vaccination with either BNT162b2 or ChAdOx1 nCoV-19, depending on availability. This sub-study included a total of 517 HCW stratified into two groups depending on SARS-CoV-2 infection prior to vaccination. 337 HCW received BNT162b2 with a 3-week dose interval (range 21-28 days), 72 HCW received BNT162b2 with a 6-week dose interval (range 39-52 days) and 108 HCW received ChAdOx1 nCoV-19 with a 12-week dose interval (range 71-92 days). Figure 1. Demographics, prior SARS-CoV-2 infection and vaccine status of the study population are presented in Table 1.

**Figure 1.**
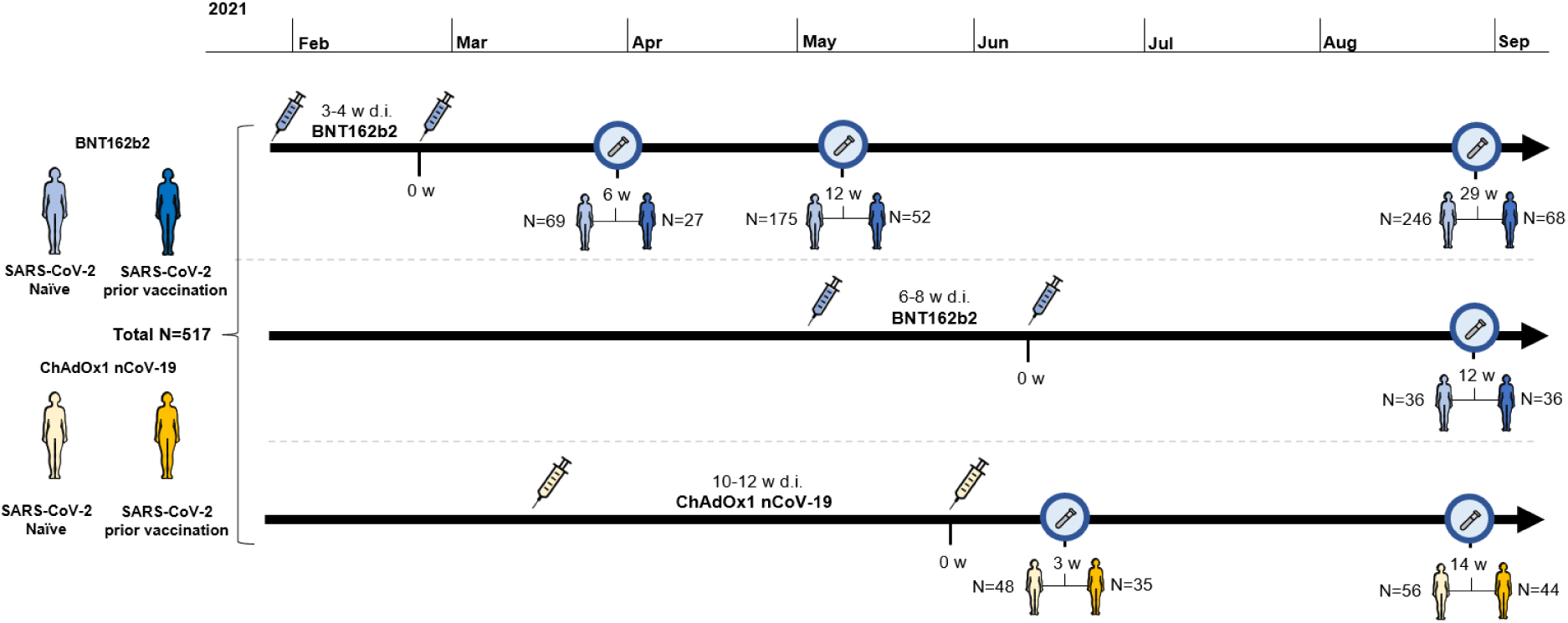
Timeline for vaccination and sample collection. The cohort is divided into participants receiving BNT162b2 with a 3-4 week and a 6–8 week dose interval and ChAdOx1 nCoV-19 with a 10-12 week dose interval. Blue characters represent vaccinees that received BNT162b2 and yellow characters represent vaccinees that received ChAdOx1 nCoV-19. Light colored characters represent SARS-CoV-2 naïve and dark colored characters represent participants with SARS-CoV-2 infection prior vaccination. Test tubes represent time for blood sampling and syringes represent time for vaccination. w; weeks. d.i.; dose interval.

**Table 1.** Demographics of study participants. Age is presented as median with IQR; inter-quartile range. Weeks post second vaccine dose refers to point in time where sampling occurred and are presented as median together with the range. d.i.; dose interval, p.v.; post second vaccine dose.

|  | SARS-CoV-2 naïve |  |  | SARS-CoV-2 recovered |  |  |
| --- | --- | --- | --- | --- | --- | --- |
|  | N | Age (IQR) | Female (%) | N | Age (IQR) | Female (%) |
| **BNT162b2 (3-4 weeks d.i.)** |  |  |  |  |  |  |
| **Total** | **262** |  |  | **75** |  |  |
| 6 weeks p.v. (5-8) | **69** | 50 (40-56) | 84 | **27** | 50 (41-55) | 81 |
| 12 weeks p.v. (11-14) | **175** | 51 (40-57) | 90 | **52** | 48 (39-56) | 81 |
| 29 weeks p.v. (27-30) | **246** | 50 (40-57) | 87 | **68** | 49 (39-57) | 82 |
| **BNT162b2 (6-8 weeks d.i.)** |  |  |  |  |  |  |
| **Total** | **36** |  |  | **36** |  |  |
| 12 weeks p.v. (11-14) | **36** | 61 (54-62) | 92 | **36** | 54 (40-60) | 86 |
| **ChAdOx1 nCoV-19 (10-12 weeks d.i.)** |  |  |  |  |  |  |
| **Total** | **61** |  |  | **47** |  |  |
| 3 weeks p.v. (2-3) | **48** | 52 (42-57) | 90 | **35** | 52 (44-58) | 86 |
| 14 weeks p.v. (13-15) | **56** | 51 (38-56) | 89 | **44** | 52 (44-57) | 86 |

### Effect of previous SARS-CoV-2 infection on antibody responses over seven months following BNT162b2 vaccination

Almost all (99.8%) participants had detectable levels of spike IgG antibodies after vaccination. Notably, at all sampling time points, spike IgG GMTs were markedly higher in previously SARS-CoV-2 infected vaccinees compared to SARS-CoV-2 naïve vaccinees (all p<0.001). Table 2 and Figure 2A. A 2-fold decrease of spike IgG GMTs between week 6 and 12, and a 6.6-fold decrease between week 6 and 29, were observed in SARS-CoV-2 naïve BNT162b2 vaccinees. For vaccinees with SARS-CoV-2 infection prior to vaccination, a 1.5-fold decrease of GMTs was observed between week 6 and 12, and a 3.6-fold decrease was observed between week 6 and 29 post vaccination.

**Table 2.**
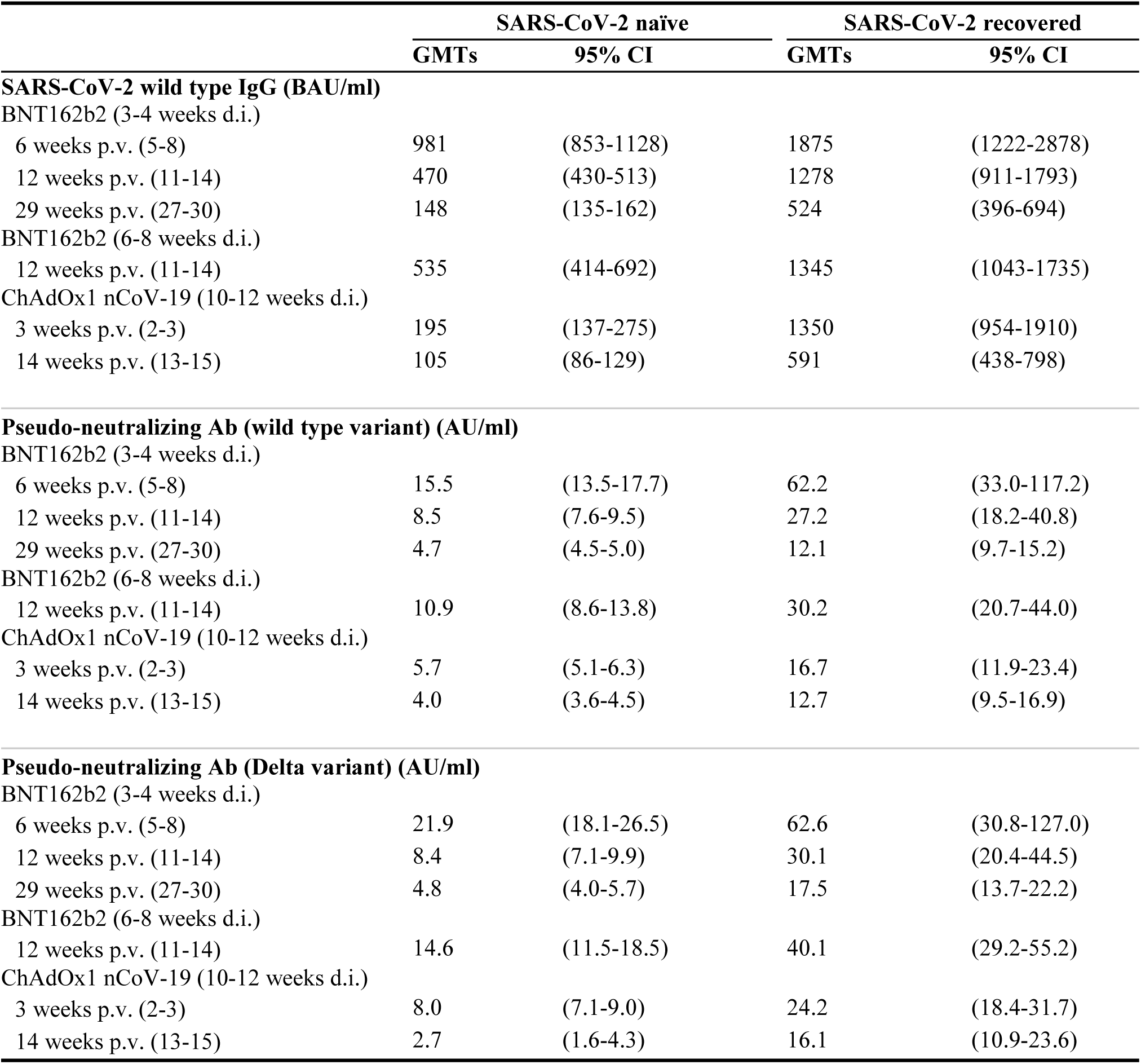
Geometric mean titers (GMTs) of SARS-CoV-2 wild type spike IgG and pseudo-neutralizing antibodies in the different study groups. Weeks post second vaccine dose refers to point in time where sampling occurred and are presented as median together with the range. CI; confidence interval, Ab; antibodies, d.i.; dose interval, p.v.; post second vaccine dose, AU; arbitrary unit, BAU; binding antibody unit.

|  | SARS-CoV-2 naïve |  | SARS-CoV-2 recovered |  |
| --- | --- | --- | --- | --- |
|  | GMTs | 95% CI | GMTs | 95% CI |
| **SARS-CoV-2 wild type IgG (BAU/ml)** |  |  |  |  |
| BNT162b2 (3-4 weeks d.i.) |  |  |  |  |
| 6 weeks p.v. (5-8) | 981 | (853-1128) | 1875 | (1222-2878) |
| 12 weeks p.v. (11-14) | 470 | (430-513) | 1278 | (911-1793) |
| 29 weeks p.v. (27-30) | 148 | (135-162) | 524 | (396-694) |
| BNT162b2 (6-8 weeks d.i.) |  |  |  |  |
| 12 weeks p.v. (11-14) | 535 | (414-692) | 1345 | (1043-1735) |
| ChAdOx1 nCoV-19 (10-12 weeks d.i.) |  |  |  |  |
| 3 weeks p.v. (2-3) | 195 | (137-275) | 1350 | (954-1910) |
| 14 weeks p.v. (13-15) | 105 | (86-129) | 591 | (438-798) |
| **Pseudo-neutralizing Ab (wild type variant) (AU/ml)** |  |  |  |  |
| BNT162b2 (3-4 weeks d.i.) |  |  |  |  |
| 6 weeks p.v. (5-8) | 15.5 | (13.5-17.7) | 62.2 | (33.0-117.2) |
| 12 weeks p.v. (11-14) | 8.5 | (7.6-9.5) | 27.2 | (18.2-40.8) |
| 29 weeks p.v. (27-30) | 4.7 | (4.5-5.0) | 12.1 | (9.7-15.2) |
| BNT162b2 (6-8 weeks d.i.) |  |  |  |  |
| 12 weeks p.v. (11-14) | 10.9 | (8.6-13.8) | 30.2 | (20.7-44.0) |
| ChAdOx1 nCoV-19 (10-12 weeks d.i.) |  |  |  |  |
| 3 weeks p.v. (2-3) | 5.7 | (5.1-6.3) | 16.7 | (11.9-23.4) |
| 14 weeks p.v. (13-15) | 4.0 | (3.6-4.5) | 12.7 | (9.5-16.9) |
| **Pseudo-neutralizing Ab (Delta variant) (AU/ml)** |  |  |  |  |
| BNT162b2 (3-4 weeks d.i.) |  |  |  |  |
| 6 weeks p.v. (5-8) | 21.9 | (18.1-26.5) | 62.6 | (30.8-127.0) |
| 12 weeks p.v. (11-14) | 8.4 | (7.1-9.9) | 30.1 | (20.4-44.5) |
| 29 weeks p.v. (27-30) | 4.8 | (4.0-5.7) | 17.5 | (13.7-22.2) |
| BNT162b2 (6-8 weeks d.i.) |  |  |  |  |
| 12 weeks p.v. (11-14) | 14.6 | (11.5-18.5) | 40.1 | (29.2-55.2) |
| ChAdOx1 nCoV-19 (10-12 weeks d.i.) |  |  |  |  |
| 3 weeks p.v. (2-3) | 8.0 | (7.1-9.0) | 24.2 | (18.4-31.7) |
| 14 weeks p.v. (13-15) | 2.7 | (1.6-4.3) | 16.1 | (10.9-23.6) |

**Figure 2.**
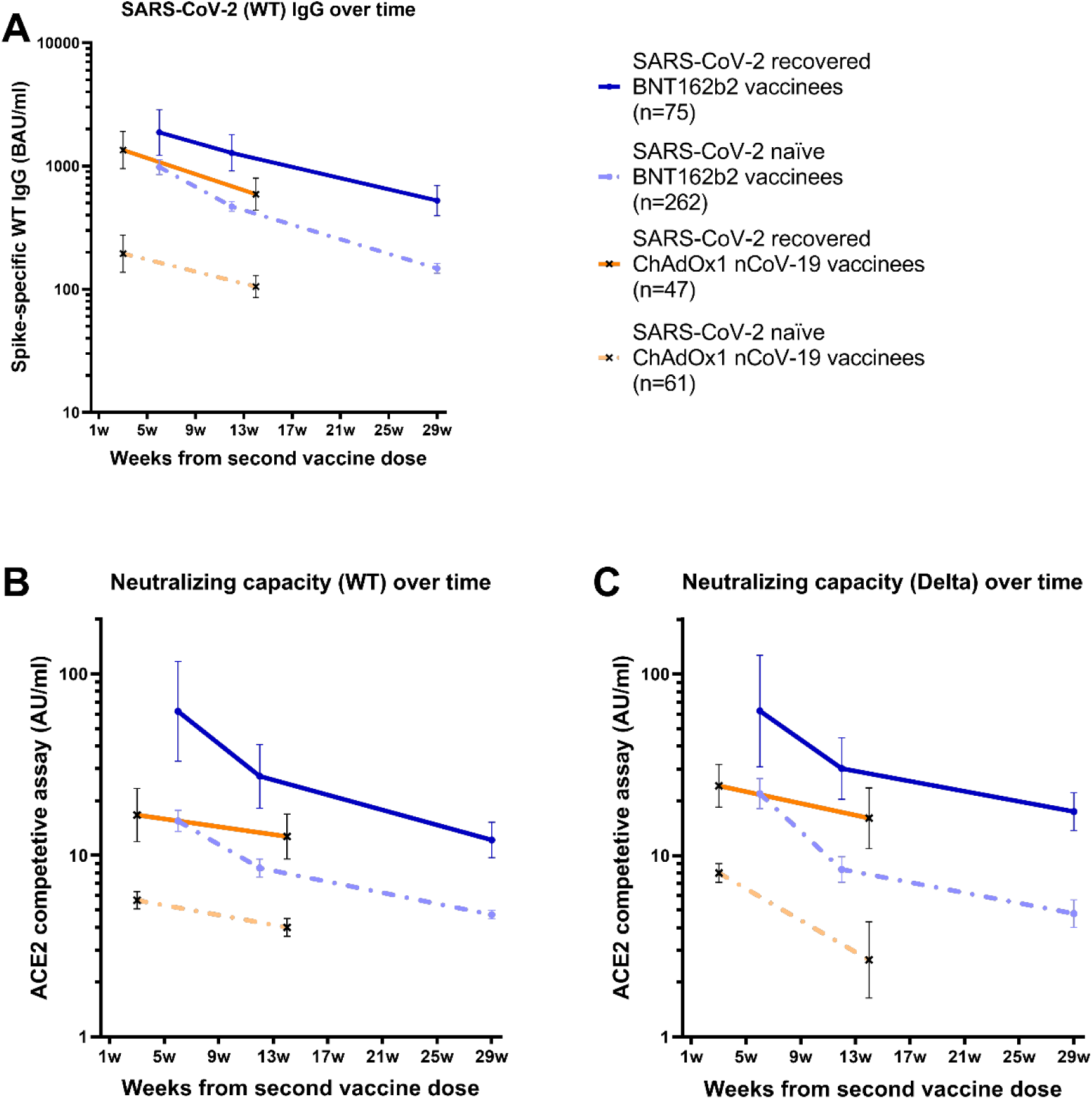
Binding and pseudo-neutralizing antibody titers over time following BNT162b2 and ChAdOx1 nCoV-19 vaccination with and without prior SARS-CoV-2 infection. A) Binding antibody titers against SARS-CoV-2 wild type over 7 months following the second BNT162b2 dose and 3 months following the second ChAdOx1 nCoV-19 dose in SARS-CoV-2 recovered and naïve vaccinees, B) pseudo-neutralizing antibodies against the wild type over 7 months following the second BNT162b2 dose and 3 months following the second ChAdOx1 nCoV-19 dose in SARS-CoV-2 recovered and naïve vaccinees, and C) pseudo-neutralizing antibodies against the Delta variant type over 7 months following the second BNT162b2 dose and 3 months following the second ChAdOx1 nCoV-19 dose in SARS-CoV-2 recovered and naïve vaccinees. Dots and crosses represent geometric mean titers and bars represent 95 % CI. Solid lines represent SARS-CoV-2 recovered vaccinees and dotted lines represent SARS-CoV-2 naïve vaccinees. WT; wild type, BAU; binding antibody units, AU; arbitrary units.

Spike IgG titers correlated strongly to pseudo-neutralizing antibody titers against both the WT and the Delta variant (Spearman’s rank correlation coefficient 0.92, and 0.83, respectively, both p<0.0001). Consistent with spike IgG, substantially higher GMTs of pseudo-neutralizing antibodies were observed in SARS-CoV-2 recovered vaccinees, as compared to SARS-CoV-2 naïve vaccinees, at all sampling time points (all p<0.001). Table 2 and Figure 2 B-C. Among SARS-CoV-2 naïve vaccinees, GMTs of pseudo-neutralizing antibodies against the WT and Delta variant decreased by 1.8-fold and 2.6-fold between week 6 and 12, and by 3.3-fold and 4.6-fold between week 6 and 29, respectively. For SARS-CoV-2 recovered vaccinees, GMTs of pseudo-neutralizing antibodies against the WT and Delta variant decreased by 2.3-fold and 2.1-fold between weeks 6 and 12, and by 5.1-fold and 3.6-fold between weeks 6 and 29, respectively. Table 2 and Figure 2B-C. Notably, pseudo-neutralizing GMTs against the Delta variant reached similar trajectories as pseudo-neutralizing GMTs against the WT, indicating a similar trend in response for the currently circulating Delta variant as for the initially circulating WT. Table 2 and Figure 2B-C.

### Effect of BNT162b2 dose interval on antibody levels

It has been reported that a longer interval between BNT162b2 doses can result in higher antibody titers *(27)*. In this cohort, BNT162b2 vaccinees were administered with a 3–4-week dose interval in January to February 2021, and with a prolonged dose interval of 6-8 weeks in April to July 2021, allowing for a comparison in immune responses in these two groups.

12 weeks post second dose, there were no significant differences in spike IgG GMTs following BNT162b2 with the 3-4 week and the 6–8-week dose interval, regardless of prior SARS-CoV-2 infection (all p>0.05). Table 2. Pseudo-neutralizing antibody GMTs against both the WT and Delta variant were, however, slightly but significantly increased 12 weeks following second dose after the prolonged dose interval (p=0.016 and p=0.017, respectively) in SARS-CoV-2 naïve vaccinees when adjusted for sex and age. A similar trend was seen in SARS-CoV-2 recovered vaccinees, although the differences were not significant (all p>0.05). Table 2.

### Effect of previous SARS-CoV-2 infection on antibody responses over three months following ChAdOx1 nCoV-19 vaccination

We next analysed the effect of previous SARS-CoV-2 infection on immune responses after vaccination with the ChAdOx1 nCoV-19 vaccine. At 12 weeks post vaccination, spike IgG GMTs in SARS-CoV-2 naïve ChAdOx1 nCoV-19 vaccinees were 4.5-fold lower compared to SARS-CoV-2 naïve BNT162b2 vaccinees (p<0.001). As observed after BNT162b2 vaccination, spike IgG GMTs were substantially increased in SARS-CoV-2 recovered ChAdOx1 nCoV-19 vaccinees compared to in SARS-CoV-2 naïve vaccinees at all sampling time points (all p<0.001). Table 2 and Figure 2A-C. In SARS-CoV-2 naïve vaccinees, spike IgG GMTs declined 1.9-fold between week 3 and 14 post vaccination, whereas it declined 2.3-fold between week 3 and 14 post vaccination in SARS-CoV-2 recovered vaccinees. Table 2 and Figure 2 A-C.

Consistent with results for binding antibodies, substantially higher GMTs of pseudo-neutralizing antibodies were observed in SARS-CoV-2 recovered ChAdOx1 nCoV-19 vaccinees compared to in SARS-CoV-2 naïve vaccinees at all sampling time points (all p<0.001). Table 2 and Figure 2 B-C. In SARS-CoV-2 naïve vaccinees, GMTs against the WT and Delta variant decreased by 1.4-fold and 3-fold between week 3 and 14, respectively, whereas they decreased by 1.3-fold and 1.5-fold, respectively, in SARS-CoV-2 recovered vaccinees. Similar to findings following BNT162b2 vaccination, GMTs against the Delta variant reached comparable levels and trajectories as GMTs against the WT. Table 2 and Figure 2 B-C.

### Live-microneutralization compared to pseudo-neutralization

As a slightly higher capacity to inhibit binding of the Delta variant compared to the WT was found using the pseudo-neutralizing assay (Figure 2B-C), a live-neutralizing assay was performed in a sub-set of 34 participants (17 SARS-CoV-2 naïve BNT162b2 vaccinees and 17 SARS-CoV-2 naïve ChAdOx1 nCoV-19 vaccinees). While results from these two assays correlated with regard to the WT and the Delta variant (rS=0.66 and 0.63 respectively, both p<0.0001), the live-microneutralization assay resulted in 25.2 % lower titers against the Delta variant compared to the WT.

### Neutralization capacity against SARS-CoV-2 variants in vaccinees with and without SARS-CoV-2 infection prior to vaccination

In light of the substantial increases in antibody titers observed in SARS-CoV-2 recovered vaccinees as compared to SARS-CoV-2 naïve vaccinees, we proceeded to determine if it also had an effect on neutralizing capacity against SARS-CoV-2 variants. Pseudo-neutralizing antibody titers were assessed against ten SARS-CoV-2 variants, including all variants of concern (VOC) (B.1.1.7 (Alpha), B.135.1 (Beta), P.1 (Gamma) and B.1.617.2 (Delta)). Notably, pseudo-neutralizing antibody titers against all ten tested SARS-CoV-2 variants were at least 2 respectively 3-fold higher in SARS-CoV-2 recovered as compared to naïve vaccinees following BNT162b2 and ChAdOx1 nCoV-19, respectively (all p<0.001). Figure 3 A-C and Table S1.

**Figure 3.**
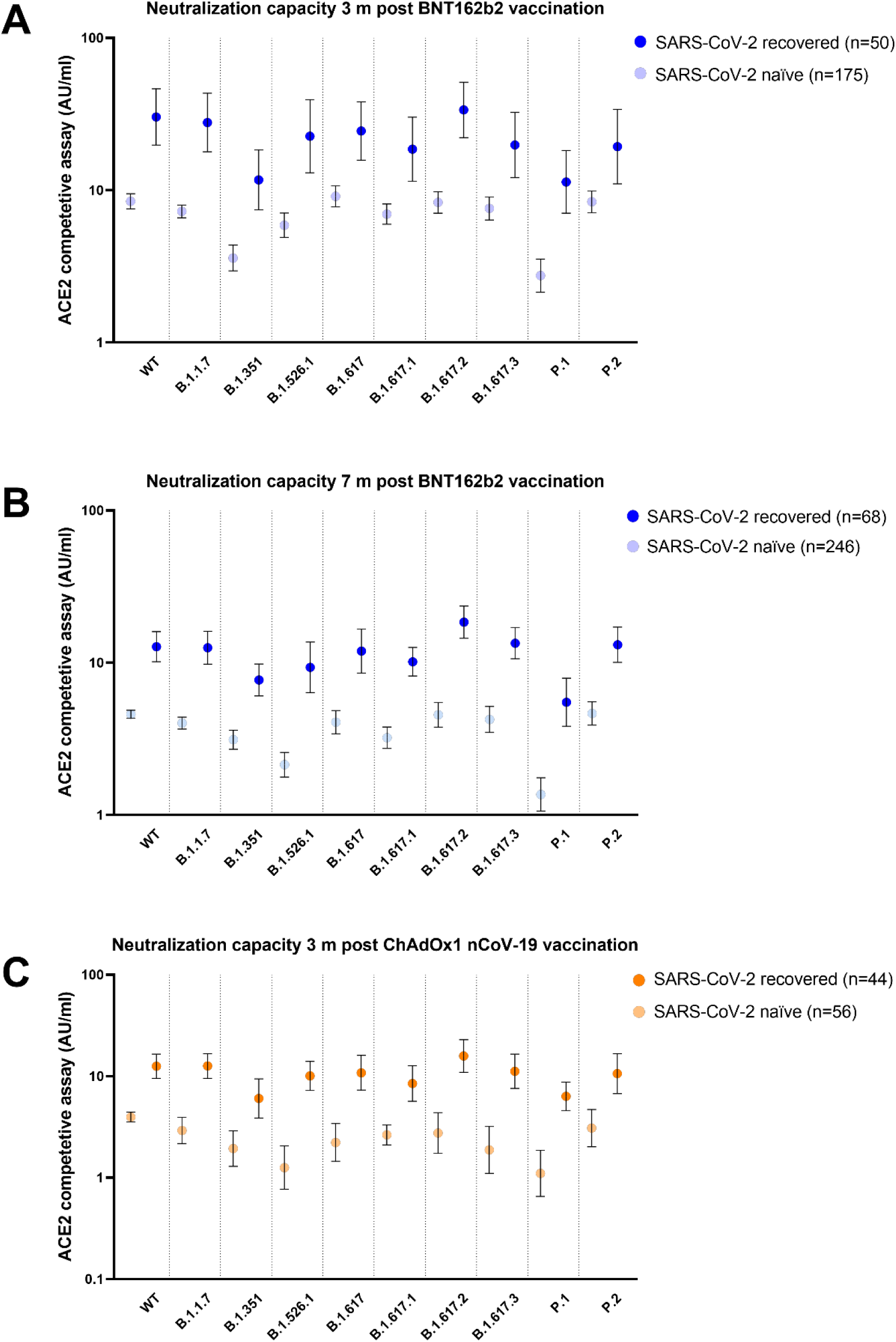
Neutralizing capacity over time. Neutralizing capacity A) 3 months and B) 7 months post BNT162b2 vaccination and C) 3 months post ChAdOx1 nCoV-19 vaccination against ten SARS-CoV-2 variants including the wild type (WT), B.1.1.7 (Alpha), B.1.351 (Beta), B.1.526.1 (New York), B.1.617 (India), B.1.617.1 (Kappa), B.1.617.2 (Delta), B.1.617.3 (India), P.1 (Gamma), and P.2 (Zeta) variants. Dots represent geometric mean titers and bars represent 95 % CI. Dark colored dots represent SARS-CoV-2 recovered vaccinees and light-colored dots represent SARS-CoV-2 naïve vaccinees.

### SARS-CoV-2 specific memory T cell responses

We next proceeded to investigate SARS-CoV-2 directed T cell responses in a subset of SARS-CoV-2 naïve and recovered vaccinees three months post ChAdOx1 nCoV-19 as well as three- and seven-months post BNT162b2. As expected, memory T-cell responses were stronger in SARS-CoV-2 recovered vaccinees compared to SARS-CoV-2 naïve vaccinees. We used a whole blood IGRA assay based on a pool of eight peptides derived from the spike protein (peptides that do not show any evidence of overlap with common cold coronaviruses and thus do not pose a risk of detecting T cell responses inflicted by a previous non-SARS-CoV-related infection *(13, 28)*. IFN-γ GMTs were more than 6-fold higher in SARS-CoV-2 recovered BNT162b2 vaccinees during the whole follow-up period of seven months (166.4 pg/ml (95% CI 98.5-281.2) and 116.8 pg/ml (95% CI 34.6-394.7) in SARS-CoV-2 recovered vaccinees three- and seven-months post BNT162b2 vs. 26.8 pg/ml (95% CI 45.5-49.6) and 18.7 pg/ml (95% CI 9.5-36.6) in SARS-CoV-2 naïve vaccinees; p<0.01). Figure 4 A-B. Similar differences were found three months post ChAdOx1 nCoV-19, with a more than 4-fold increase in IFN-γ geometric mean levels in SARS-CoV-2 recovered vaccinees (38.0 pg/ml (95% CI 20.4-70.9) in SARS-CoV-2 recovered vaccinees vs. 8.0 pg/ml (95% CI 3.5-18.5) in SARS-CoV-2 naïve vaccinees; p<0.01). Figure 4 C.

**Figure 4.**
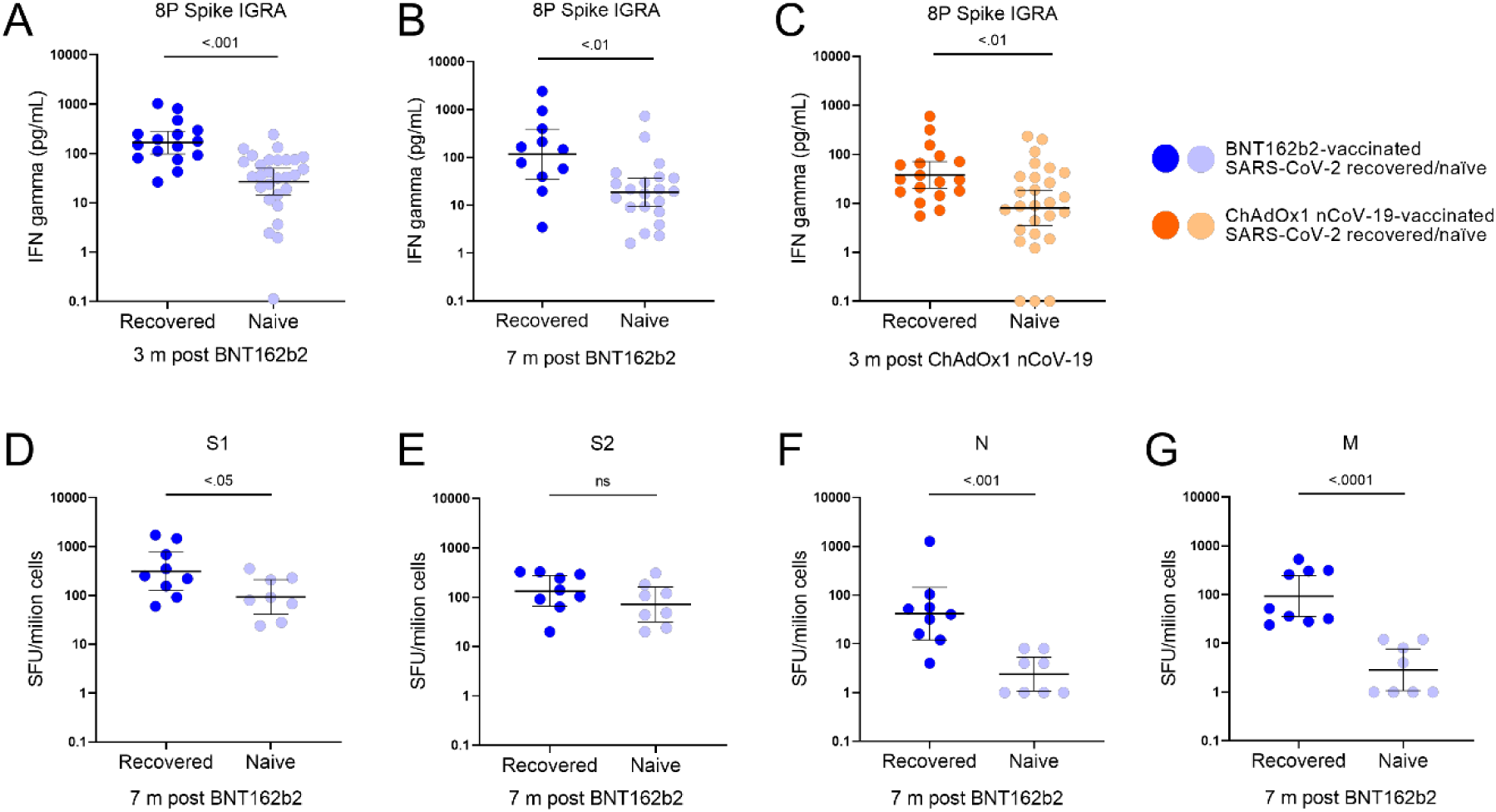
Memory T cell responses three and seven months following BNT612b2 and ChAdOx1 nCoV-19 vaccination in SARS-CoV-2 recovered and naïve participants. A-C; Whole blood IGRA assay using a SARS-CoV-2-specific peptide pool (8 SARS-CoV-2-specific peptides covering the SARS-CoV-2 spike protein *(13, 28)*). D-G: T-cell responses against S1, S2, N and M analyzed against T-SPOT® Discovery SARS-CoV-2 kit (Oxford Immunotec). Blue dots represent BNT612b2 vaccinees (A-B, D-G), and orange dots represent ChAdOx1 nCoV-19 vaccinees (C). Dark-colored dots represent SARS-CoV-2 recovered participants, light-colored dots represent SARS-CoV-2 naïve. Lines represent geometric mean and error bars represent 95% confidence interval.

These findings were corroborated using a commercial T-spot assay, at seven months post BNT162b2 vaccination, with a set of peptide pools that cover the S1, S2, N, and M protein. With this assay we observed more than a 4-fold increased response against the S1 protein in SARS-CoV-2 recovered vaccinees as compared to in SARS-CoV-2 naïve vaccinees (556 SFU/million cells in SARS-CoV-2 recovered vs. 135 SFU/million cells in SARS-CoV-2 naïve vaccinees; p<0.05). Figure 4 D-G. No significant difference in response against the S2 protein was observed between the two groups. Notably, responses against the N and M peptide pools remained enhanced in SARS-CoV-2 recovered vaccinees, implying long lasting infection acquired immune responses against these proteins, but also confirming the serological status prior vaccination.

## Discussion

Using a large cohort with longitudinally collected blood samples, we show that vaccination following SARS-CoV-2 infection resulted in remarkably and sustained enhancement of both humoral and cellular immune responses, with an increased neutralizing potency and breadth against SARS-CoV-2 variants as compared to vaccination in SARS-CoV-2 naïve individuals. Furthermore, we reveal a significant decline in BAU/ml and pseudo-neutralizing antibody titers over the first months following both ChAdOx1 nCoV-19 and BNT162b2 vaccination. Direct comparisons showed substantially lower titers following immunization with ChAdOx1 nCoV-19 compared to BNT162b2. These findings are important and can be of support in the decisions on booster doses and design of current and future SARS-CoV-2 vaccine programs.

We and others have previously shown that COVID-19 recovered vaccinees mount strong immune responses following mRNA *(9, 19-23)* and adenovirus-vectored vaccines *(24)*. We here extend these findings by showing that both T-cell responses (herein measured both by a traditional ELISpot assay using overlapping peptides spanning whole protein sequences as well as a whole blood IGRA based method using carefully selected SARS-CoV-2 unique S-derived peptides) and antibody titers remain substantially increased over three months following ChAdOx1 nCoV-19 and seven months following BNT162b2 in HCW with mild SARS-CoV-2 infection prior to vaccination. This suggests that the effect of a previous infection followed by vaccination on immune responses is not a temporary phenomenon. The memory compartment continues to evolve after both natural infection and vaccination, but with a lesser increase in breadth following vaccination as compared to infection *(18)*. Importantly, we found that SARS-CoV-2 recovered vaccinees developed higher neutralizing antibody responses against SARS-CoV-2 variants than SARS-CoV-2 naïve vaccinees throughout the study period. These findings suggest that vaccination of immune competent SARS-CoV-2 recovered individuals evokes improved cellular responses and antibody levels than in SARS-CoV-2 naïve vaccinees.

Our findings of substantial reductions in antibody titers over the first three months following ChAdOx1 nCoV-19 and seven months following BNT162b2 are in line with several reports of waning vaccine efficacy from countries including Israel *(4, 5)*, UK *(6, 7)* and the US *(8)*. The comparatively lower titers following ChAdOx1 nCoV-19 are also in line with prior data on lower vaccine efficacy of adenovirus-vectored vaccines as compared to mRNA vaccines *(2, 29)*. Specific antibody levels are used as a correlate of protection against infection following several established vaccines *(30)*, and for SARS-CoV-2, binding and pseudo-neutralizing antibody titers correlate inversely with the risk of infection *(14, 31)*. Recently, an 80% vaccine efficacy against symptomatic SARS-CoV-2 infection was observed at 264 BAU/ml, declining to 60% vaccine efficacy at 54 BAU/ml *(32)*, with similar results in another study *(33)*. Our findings suggest that antibody titers have declined below these thresholds in a portion of study participants within 3-7 months from the second vaccine dose, indicating a need for a third booster in a subpopulation of the vaccinated individuals Importantly, the estimations were derived in eras with a dominance of the Alpha variant, and the estimated titers may provide lower efficacy against infection with the Delta variant *(34-36)*, which seems to have a higher transmissibility *(37, 38)*.

Optimal dose interval has been proposed to convey better vaccine efficacy via improved memory formation *(17)*. A longer interval may indeed generate an improved memory formation, albeit at the price of a suboptimal protection during the longer interval between first and second dose in SARS-CoV-2 naïve populations. Although no significant differences in binding antibody titers was observed, a slight difference in pseudo-neutralization titers were found between the 3-4 week and 6–8-week dose interval in SARS-CoV-2 naïve participants, possibly indicating improved memory formation in the latter group *(39)*. Continued studies are needed to address potential differences in longevity of the immunological memory induced by different prime boost intervals and combinations of various vaccine platforms, as well as the optimal timing of additional vaccine doses.

This study is limited by the observational and single-center nature. The study cohort moreover comprised HCW, and a majority of women of general working age. Antibody trajectories may differ in older populations, and in settings without repeated viral encounters with potential boosting of the immune memory.

In summary, the striking and sustained enhanced cellular immune responses, antibody titers and neutralizing breadth in previously SARS-CoV-2 infected vaccinees as compared to SARS-CoV-2 naïve vaccinees highlights the strong impact of infection prior to vaccination. These findings suggest that prior SARS-CoV-2 infection should be taken into consideration in vaccine policy making, when planning booster doses, and in the design of current and future SARS-CoV-2 vaccine programs.

## Data Availability

The anonymized datasets generated during and/or analysed during the current study are available from the corresponding author on reasonable request.

## Funding

Jonas & Christina af Jochnick foundation (CT)

Lundblad family foundation (CT)

Region Stockholm (CT)

Knut and Alice Wallenberg foundation (CT, SM)

Jonas Söderquist’s scholarship (CT)

Science for Life Laboratory (PN)

Erling-Persson family foundation (SoH)

Center for Innovative Medicine (JK)

Swedish Research Council (JK)

## Author contributions

Conceptualization: SeH, UM, MÅ, CT, SM

Methodology: SeH, UM, WC, MG, SM, JK MÅ, CT

Investigation: SeH, UM, NGN, HN, WC, KB, JK, PN, SoH, SM, MÅ, CT

Visualization: SeH, MG

Funding acquisition: CT, PN, SoH, JK, SM

Project administration: NGN, SeH, MÅ, CT

Supervision: CT, MÅ, SoH, MP, SM

Writing – original draft: SH, CT

Writing – review & editing: All authors

## Supplementary material

### Materials and methods

#### Study population

The participants of the COMMUNITY study are followed every four months since inclusion in April 2020, where SARS-CoV-2 spike-specific IgG are analyzed by multiplex antigen bead array *(40)*. Previous SARS-CoV-2 infection was confirmed by seroconversion at any of the follow-up visits before vaccination. Date and type of vaccine was obtained through the Swedish vaccination register (VAL Vaccinera). The study is approved by the Swedish Ethical Review Authority (dnr 2020-01653) and written informed consent was obtained from all study participants.

#### Binding and pseudo-neutralizing antibodies

Binding antibodies (IgG) and pseudo-neutralizing antibodies against SARS-CoV-2 wild type B.1.1.7 (Alpha), B.1.351 (Beta), B.1.526.1 (New York), B.1.617 (India), B.1.617.1 (Kappa), B.1.617.2 (Delta), B.1.617.3 (India), P.1 (Gamma), and P.2 (Zeta) variants, were measured in post vaccination samples using the V-PLEX SARS-CoV-2 Panel 13 (Meso Scale Diagnostics, USA) for IgG and ACE2 (quantifying the ability to inhibit the binding between the ACE2 receptor and the spike protein), respectively. Binding antibody titers for WT were calibrated against the WHO standard *(16)* according to the manufacturer’s instructions and presented as Binding Antibody Units (BAU)/ml. Pseudo-neutralizing antibodies are expressed as arbitrary units (AU)/ml.

#### Microneutralization assay

Microneutralization based on cytopathic effects (CPE) was performed essentially as previously described *(41)*. Briefly, serum was 3-fold serially diluted, mixed with virus, incubated for 1 hour and finally added, in duplicates, to confluent Vero E6 cells in 96-well plates. Original SARS-CoV-2 WT (isolated from a Swedish patient) and the Delta variant (from Statens Serum Institut, Copenhagen, Denmark), were used. After 5 days incubation, the wells were inspected for signs of CPE by optical microscopy. Each well was scored as either neutralizing (if no signs of CPE was observed) or non-neutralizing (if any CPE was observed). The arithmetic mean neutralization titer of the reciprocals of the highest neutralizing dilutions from the two duplicates for each sample was then calculated.

#### SARS-CoV-2 specific memory T cell response

Whole blood Interferon-Gamma (IFN-γ) Release Assay (IGRA) was performed as previously described *(13, 28)*. A SARS-CoV-2-specific peptide pool was generated using 8 SARS-CoV-2-specific peptides covering the SARS-CoV-2 spike. Peptides were synthesized with a purity of >95% and contained no more than 5-mer length overlap with endemic coronaviruses *(13, 28)*. Peripheral blood was collected in lithium heparin tubes and 0.5 ml was added to glucose (2 mg/ml whole blood) and 0.9% NaCl with and without the stimulant. Samples were incubated at 37°C with 5% CO2 for 20 h. IFN-γ was analyzed in plasma using Mesoscale Discovery V-plex kit (Meso Scale Diagnostics, Maryland, USA). The whole blood IGRA was performed in a subset of 44 BNT162b2 vaccinated 3 months post vaccination (28 SARS-CoV-2 naïve, age 53 [IQR 38-63], 79% female and 16 SARS-CoV-2 recovered, age 47 [IQR 39-55], 69% female), 32 BNT162b2 vaccinated 7 months post vaccination (21 SARS-CoV-2 naïve, age 53 [IQR 43-60], 95% female and 11 SARS-CoV-2 recovered, age 51 [IQR 39-56], 82% female) and 45 ChAdOx1 nCoV-19 vaccinated participants 3 months post vaccination (27 SARS-CoV-2 naïve, age 55 [IQR 46-61], 85% female and 18 recovered, age 52 [IQR 46-60], 83% female).

Peripheral blood mononuclear cells (PBMCs) were isolated from whole blood using CPT tubes. After quantification and dilution of recovered cells, 250,000 PBMCs were plated into each well of a T-SPOT® Discovery SARS-CoV-2 kit (Oxford Immunotec), according to manufacturer’s instructions. The kit contains overlapping peptide pools covering protein sequences of six SARS-CoV-2 antigens, without HLA restriction. Peptide sequences with high homology to endemic coronaviruses were removed, but sequences that may have homology to SARS-CoV-1 were retained. Cells were incubated 20 hours and interferon-γ secreting T cells detected. The T-spot analyses were performed in a sub-set of 17 BNT162b2 vaccinees seven months post vaccination (8 SARS-CoV-2 naïve, age 58 [IQR 52-62], 100% female and 9 recovered, age 51 [IQR 35-59], 67% female).

#### Statistical analyses

Spike IgG and pseudo-neutralizing antibody titers are presented as geometric mean titers (GMTs) with 95% confidence interval (CI). Linear regression was used to compare continuous variables between groups and were adjusted for sex and age. A t test on logarithmized values was used to compare differences between the T cell analyses groups, due to small number of study participants. Multiple samples per subject were analyzed using linear-mixed effects regression with random intercepts per subject using the same adjustments as for linear regression. The correlation coefficient was calculated using Spearman’s correlation analysis. Linear regression and mixed effects regressions were performed in R (version 4.1.1) with nlme-package version 3.1.152 and contrast-package 0.22 while the remainder were done in GraphPad Prism (version 9.1.1).

