## Supplemental Table 1 for "Impact of SARS-CoV-2 infection on vaccine-induced immune responses over time" (from another version of this preprint: version 1, 2021-10-19)

|  | **Ratio (95 % CI)** | **P-value** |
| --- | --- | --- |
| **BNT612b2 (3-4 weeks d.i.)** | | |
| **12 weeks p.v.** | | |
| Wild type | 3.2 (2.1-4.7) | < 0.0001 |
| Alpha (B.1.1.7) | 3.3 (2.3-5.0) | < 0.0001 |
| Beta (B.1.351) | 2.9 (1.9-4.2) | < 0.0001 |
| Gamma (P.1) | 3.6 (2.4-5.4) | < 0.0001 |
| Delta (B.1.617.2) | 3.5 (2.4-5.2) | < 0.0001 |
| B.1.526.1 | 3.3 (2.2-4.9) | < 0.0001 |
| B.1.617 | 2.4 (1.6-3.5) | < 0.0001 |
| B.1.617.1 | 2.3 (1.6-3.5) | < 0.0001 |
| B.1.617.3 | 2.2 (1.5-3.3) | < 0.0001 |
| P.2 | 2.0 (1.4-3.0) | 0.0004 |
| **BNT612b2 (3-4 weeks d.i.)** | | |
| **29 weeks p.v.** | | |
| Wild type | 2.6 (1.8-3.6) | < 0.0001 |
| Alpha (B.1.1.7) | 2.9 (2.1-4.0) | < 0.0001 |
| Beta (B.1.351) | 2.3 (1.7-3.2) | < 0.0001 |
| Gamma (P.1) | 3.7 (2.6-5.1) | < 0.0001 |
| Delta (B.1.617.2) | 3.6 (2.6-5.1) | < 0.0001 |
| B.1.526.1 | 3.8 (2.7-5.3) | < 0.0001 |
| B.1.617 | 2.8 (2.0-3.9) | < 0.0001 |
| B.1.617.1 | 3.0 (2.1-4.1) | < 0.0001 |
| B.1.617.3 | 2.9 (2.1-4.1) | < 0.0001 |
| P.2 | 2.7 (1.9-3.8) | < 0.0001 |
| **ChAdOx1 nCoV-19 (10-12 d.i.)** | | |
| **14 weeks p.v.** | | |
| Wild type | 3.2 (1.8-5.6) | 0.0001 |
| Alpha (B.1.1.7) | 4.0 (2.3-7.0) | < 0.0001 |
| Beta (B.1.351) | 3.1 (1.8-5.5) | 0.0001 |
| Gamma (P.1) | 5.5 (3.1-9.7) | < 0.0001 |
| Delta (B.1.617.2) | 6.0 (3.4-10.6) | < 0.0001 |
| B.1.526.1 | 7.7 (4.4-13.6) | < 0.0001 |
| B.1.617 | 5.0 (2.8-8.8) | < 0.0001 |
| B.1.617.1 | 3.2 (1.8-5.7) | < 0.0001 |
| B.1.617.3 | 6.3 (3.6-11.0) | < 0.0001 |
| P.2 | 3.5 (2.0-6.2) | < 0.0001 |

Table S1. Multiplicative effect of mean pseudo-neutralization titers against ten SARS-CoV-2 variants in SARS-CoV-2 recovered vaccinees as compared to SARS-CoV-2 naïve vaccinees at 12 and 29 weeks post second vaccine dose. CI; confidence interval, d.i.; dose interval, p.v.;post second vaccine dose
